# Diabetic retinopathy screening using a portable retinal camera in Vanuatu

**DOI:** 10.1101/2022.05.25.22275597

**Authors:** Juan Caceres, Yibing Zhang, Lawrence Boe, Yunshu Zhou, Cagri Besirli, Yannis M Paulus, Julie M. Rosenthal

**Affiliations:** University of Michigan Medical School, University of Michigan, Ann Arbor, USA; Penama Provincial Health, Godden Memorial Hospital, Ambae, Vanuatu; Department of Ophthalmology and Visual Sciences, University of Michigan, Ann Arbor, MI, USA; Department of Biomedical Engineering, University of Michigan, Ann Arbor, MI, USA

**Keywords:** diabetic retinopathy, retinal screening, Vanuatu, portable retinal camera, Pictor

## Abstract

**Background and Objective:** Proof-of-concept study to test the feasibility of using an all-in-one portable retinal camera for the screening of diabetic retinopathy in the Pacific island of Vanuatu.

**Study Design/Materials and Methods:** From February 10, 2020, through February 28, 2020, 49 patients with diabetes mellitus from three islands in Vanuatu were recruited to participate in the study. Demographics, basic health data and retinal photography were obtained. A non-mydriatic, handheld camera was used (Volk Pictor Plus).

**Results:** Eleven participants (24%) had referral-warranted diabetic retinopathy. There was moderately high inter-rater reliability for our dependent variables: referral status (κ = 0.62, 95% CI 0.42-0.83), retinopathy severity (κ = 0.76, 95% CI 0.55-0.96), and clinically significant macular edema (κ = 0.50, 95% CI 0.25-0.74)

**Conclusions:** Our study confirms that portable handheld cameras can be used to obtain retinal images of sufficient quality for diabetic retinopathy screening even in resource limited environments like Vanuatu. Among this cohort, a relatively high (24%) prevalence of referral-warranted diabetic retinopathy was found in Vanuatu.

## INTRODUCTION

Diabetes mellitus is a leading cause of death and disability worldwide. Among its sequelae, diabetic retinopathy is the most common microvascular complication and affects most patients with a history of diabetes for 15 years or more. Diabetes is a worldwide epidemic and has particularly affected the islands of Vanuatu, where 10.9% of adults carry the diagnosis(1). Diabetic retinopathy is one of the major causes of visual impairment in the South Pacific Islands and the second most common diabetes-related complication in Vanuatu(2,3). Smith et al. reported a prevalence of diabetic retinopathy of 52.9% in diabetic patients studied in 2007 and emphasized the need for a “baseline screening program with ophthalmologic assessment and follow up” in order to enable adequate intervention and treatment(4). Recently, a report on Vanuatu’s non-communicable disease strategic plan, as part of the country’s government-funded healthcare system, listed annual screening for diabetic retinopathy as very cost effective(5).

Nevertheless, there is still no screening program for diabetic retinopathy in Vanuatu. Like many Pacific Island nations, Vanuatu has relied heavily on visiting teams of eye doctors to perform screenings, leaving them without a reliable and consistent screening or treatment program. However, in recent years, there has been a drastic improvement in eye care infrastructure, including the construction of a new ophthalmology clinic, the arrival of the country’s first permanent ophthalmologist, and the successful training of eye nurses in high resolution retinal photography. As such, there are now more opportunities to create an effective screening program for diabetic retinopathy. This can be done through teleophthalmology, which is among the most efficient screening strategies for diabetic retinopathy due to the cost-effectiveness and the ability to be used in both rural and urban settings(6,7).

There has been an increase in the accessibility of portable, high resolution retinal fundus cameras worldwide, which have been shown to have high-sensitivity for referral-warranted diabetic retinopathy(8–10). Due to their portability, relatively low cost, ease of use, and wide availability, these retinal cameras have the potential to be transported to the most remote islands of Vanuatu to aid in nationwide screening (11).

As such, the objective of this proof-of-concept study is to test the feasibility of using an all-in-one portable, handheld retinal camera for the screening of referral-warranted diabetic retinopathy. We also aim to better understand the prevalence of diabetic retinopathy in a cohort of patients with diabetes in three islands of Vanuatu and evaluate the effect of demographic and lifestyle factors on the presence or absence of referral-warranted diabetic retinopathy in these patients with diabetes.

## METHODS

This study was approved by the Institutional Review Board at the University of Michigan (HUM00171292, Diabetic Retinopathy in Vanuatu) and the Ethics Committee at the Vanuatu Ministry of Health. Informed consent was obtained from all individuals participating in the described study.

Study participant inclusion criteria included patients aged 18 years or older with Type 1 or Type 2 diabetes who had an appointment at the non-communicable disease clinic of the three principal hospitals (Vila Central, Godden Memorial, and Northern Provincial Hospital) on the Vanuatu islands of Efate, Ambae, and Santo, respectively. Patients were recruited and data was obtained from February 10, 2020, through February 28, 2020. Patients were excluded if they were under 18-years old, if they declined participation, or if they could not consent to the study. There were 49 participants included in the study. Additionally, three patients were excluded from retinal image analysis due the lack of gradable images.

### Procedures

Survey data was obtained by two medical students (JC and YZ) from the University of Michigan with the aid of a nurse language interpreter. Copies of the survey and consent forms were also provided both in English and Bislama as desired. Participants were asked to complete a survey that included demographic factors (including age and sex), diabetic self-management, and lifestyle factors. Visual acuity, point-of care fasting blood sugar levels (mmol/L), blood pressure, and body mass index (BMI) were also obtained from the participant if available.

Retinal photography was taken in a darkened room using a non-mydriatic handheld portable fundus digital imaging camera system, the Volk Pictor Plus (Volk Optical, Mentor, OH), which takes 40 degree field of view fundus images. Tropicamide 1% ophthalmic was used for pupil dilation to facilitate image acquisition if patients consented to do so. Participants were not required to use dilating drops for the study, but when consent was obtained, drops were instilled approximately ten minutes prior to retinal photography. Non-mydriatic retinal images were obtained in ten participants (20.4%) who declined dilating eye drops. Mydriatic retinal images were obtained in the remaining thirty-nine participants (79.6%). Three patients were excluded from the analysis of referral status because gradable retinal images were not obtained. Photographs were taken by medical students (JC and YZ) with about two hours of hands-on practice with the Volk Pictor Plus in addition reviewing the operating manual. The photographer captured three retinal images of the macula, fovea, optic disc, vascular arcades, and adjacent structures to ensure adequate field of view. The gold-standard photographic screening technique for diabetic retinopathy is seven-field, 30 degree mydriatic tabletop retinal photography comprising the posterior 90 degrees of the retina(12). Subsequent studies have demonstrated that three-field, 45 degree nonmydriatic fundus photography was effective in grading DR to determine referral-warranted disease, whereas a single 45 degree photograph was insufficient to accurately grade DR(13–15). Given that the Pictor Plus field of view is approximately 40 degrees, three photos were acquired per eye in this study to determine referral-warranted status.

Images were graded for image quality (excellent, acceptable, and not gradable) based on a previously published scale(10). A photograph was considered excellent if it was in focus and the entire posterior pole was visualized. An image was considered acceptable if it was overexposed, underexposed, or out of focus but adequate to determine the presence or absence of pathology. An ungradable photograph was one in which the image was out of focus or obscured without being able to determine pathology. This grading scheme was adapted from the National Health Service (United Kingdom) Diabetic Grading Forms(16–18). Images were stored in the camera’s internal memory and linked with the corresponding survey data using a de-identified number.

### Remote Image Interpretation

After data acquisition, images were exported to a HIPAA secure online database and stored as individual JPEG images. Images were grouped by eye (OD, OS). A total of 209 images were taken with the Pictor plus. Images and subfolders were given a random identifier in order to mask the two retina specialists (YMP and JR). Prior to initiation of image grading, there was 100% inter-rater agreement for referral-warranted diabetic retinopathy on a sample of 20 randomly selected images.

Images were graded based on image quality, the presence or absence of diabetic retinopathy, and the severity level of the retinopathy by two masked retina specialists (YMP and JR). Grading was based on the international clinical diabetic retinopathy and diabetic macular edema disease severity scales(19). Clinically significant macular edema was defined as retinal thickening or hard exudates approaching the center of the macula or involving the center of the macula (within 500 microns)(20). Moreover, image quality was graded based on gradeability (see above). Retinopathy severity level was evaluated as ungradable, no retinopathy, mild non-proliferative diabetic retinopathy (NPDR), moderate NPDR, severe NPDR, and proliferative diabetic retinopathy (PDR). Clinically significant macular edema (CSME) was also graded as present, absent, or ungradable. Any patient with CSME or with a retinopathy severity level greater than mild NPDR was considered referral-warranted (Figure 1). If CSME or retinopathy was ungradable, the image was considered ungradable. Discrepancies (gradable vs. ungradable) in grades between readers remained de-identified and were reviewed a second time. Any further discrepancies were adjudicated by a third retina specialist grader (CB).

**Figure 1.**
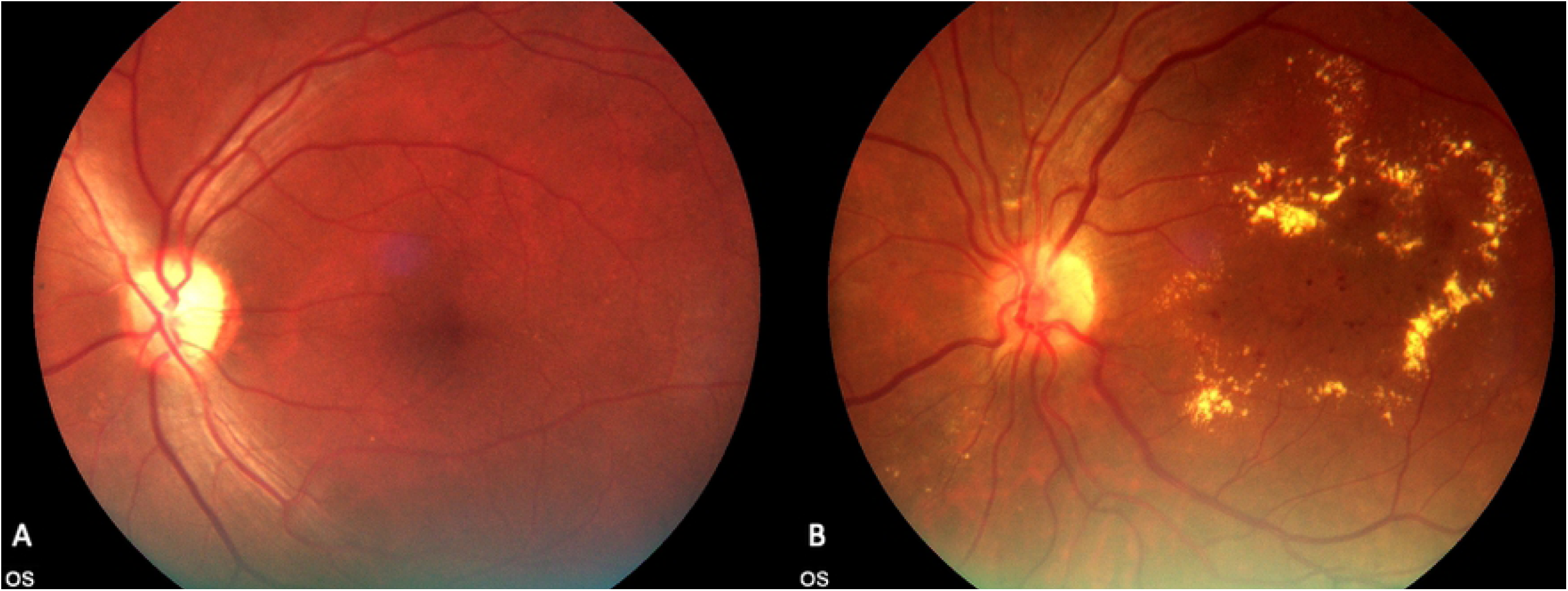
Gradable retinal images taken during the study; (A) no diabetic retinopathy; (B) referral-warranted diabetic retinopathy with clinically-significant macular edema.

### Statistical Analysis

Survey data, visual acuity, fasting blood sugar levels (mmol/L), blood pressure, and BMI were summarized both by location and by referral status. For referral status, only gradable images were used. Participants were included in the analysis if there was at least one eye that was gradable. Descriptive statistics were performed on retinal images based on grading. Either t-test or Wilcoxon rank sum test for continuous variables were used depending on the distribution of the data; and either chi-square test or Fisher’s exact tests were used for categorical variables depending on the distribution of the data. These tests were performed using SAS version 9.4 (SAS Institute, Cary, NC, USA). Inter-observer reliability was assessed for quality of retinal images, retinopathy severity, CSME, and referral status using Cohen’s kappa statistic(21). Kappa statistic was performed using the Graphpad Quickcalcs website (www.graphpad.com/quickcalcs/kappa1, San Diego, CA).

## RESULTS

Forty-nine patients representing the islands of Efate, Santo, and Ambae were included in the study. Study participants were divided equally by sex and had a mean (± standard deviation) age of 57±10. Body mass index (BMI, 27±4kg/m^2^), systolic blood pressure (138±22 mmHg), and blood glucose levels (14±5.8 mmol/L) were also similar between islands. Most participants received their diabetes diagnoses during routine screening (86%) and reported adherence to their diabetes medication regimen (84%). About half of study participants reported a dilated eye exam in the past (Table 1). More than half of participants reported having vision (65%) or foot (63%) complications as a result of their diabetes. There were 11 participants (24%) with referral-warranted diabetic retinopathy versus 35 participants (76%) who did not have referral-warranted diabetic retinopathy (Table 2). There were no significant differences in age or sex between referral and non-referral-warranted patients. BMI (27.7 vs 27.2 kg/m^2^), systolic blood pressure (138.3 vs 138.0 mm Hg), blood glucose levels (12.8 vs 13.9 mmol/L), and visual acuity were also similar between referral and non-referral-warranted groups (all p > 0.05). Most patients in both groups (81.8% vs 82.8%) reported adequate adherence to their diabetic medication regimen.

**Table 1.** Demographics and diabetes management by island in Vanuatu

| **Variable** | **Total**<br>**(n = 49)** | **Port Vila**<br>**(n = 23)** | **Santo**<br>**(n = 19)** | **Ambae**<br>**(n = 7)** |
| --- | --- | --- | --- | --- |
| Age (years; mean, SD) | 57 (10) | 57 (10) | 56 (10) | 59 (13) |
| Sex (male, %) | 23 (47) | 7 (30) | 14 (74) | 2 (29) |
| BMI (mean, SD) | 27 (4.0) | 29 (2.9) | 26 (3.9) | 25 (5.1) |
| Blood Pressure (mmHg) |  |  |  |  |
| Systolic (mean, SD) | 138 (22) | 138 (23) | 139 (26) | 139 (9.3) |
| Diastolic (mean, SD) | 83 (13) | 80 (11) | 86 (14) | 83 (11) |
| Blood glucose<sup>a</sup> (mmol/L; mean, SD) | 14 (5.8) | 14 (4.8) | 13 (6.9) | 12 (5.7) |
| Visual Acuity |  |  |  |  |
| LogMAR (mean, SD) |  |  |  |  |
| OD | 0.23 (0.24) | 0.25 (0.24) | 0.17 (0.27) | 0.32 (0.19) |
| OS | 0.23 (0.23) | 0.26 (0.24) | 0.19 (0.26) | 0.24 (0.11) |
| Snellen Equivalent (mean, SD) |  |  |  |  |
| OD | 20/34 (35) | 20/36 (35) | 20/30 (37) | 20/42 (31) |
| OS | 20/34 (34) | 20/36 (35) | 20/31 (36) | 20/35 (26) |
| Location of Diabetes Diagnosis (%) |  |  |  |  |
| During routine screening | 42 (86) | 20 (87) | 16 (84) | 6 (86) |
| During hospitalization | 7 (14) | 3 (13) | 3 (16) | 1 (14) |
| Prior dilated eye exam (%) | 27 (55) | 12 (52) | 10 (53) | 5 (71) |
| Reported adherence to diabetic medication regimen (%) |  |  |  |  |
| Somewhat | 5 (10) | 3 (13) | 2 (11) | 0 (0) |
| Very much | 41 (84) | 19 (83) | 15 (79) | 7 (100) |
| Unknown | 3 (6.1) | 1 (4.3) | 2 (11) | 0 (0) |

<sup>a</sup> Point of care blood glucose (normal 4.0-5.4 mmol/L)
Abbreviations: SD, standard deviation; BMI, body mass index; LogMAR, Logarithm of the Minimum
Angle of Resolution; OD, right eye; OS, left eye.

**Table 2.** Demographics and diabetes management by diabetic retinopathy referral status ^a^

| Variable (n = 46) | Referral<br>n = 11 | Non-referral<br>n = 35 | p-value |
| --- | --- | --- | --- |
| Age (years; mean, SD) | 55.5 (8.8) | 57.4 (10.3) | .2020 |
| Sex (male) | 5 (45.5) | 18 (51.4) | .7296 |
| BMI (mean, SD) | 27.7 (3.6) | 27.2 (3.9) | .7189 |
| Blood Pressure (mmHg) |  |  |  |
| Systolic (mean, SD) | 138.3 (33.6) | 138.0 (18.8) | .6317 |
| Diastolic (mean, SD) | 84.4 (17.7) | 81.4 (10.9) | .8793 |
| <sup>b</sup>Blood glucose (mmol/L; mean, SD) | 12.8 (5.3) | 13.9 (6.0) | .7409 |
| Visual Acuity |  |  |  |
| LogMAR (mean, SD) |  |  |  |
| OD | 0.22 (0.20) | 0.21 (0.25) | .6546 |
| OS | 0.12 (0.17) | 0.24 (0.24) | .1706 |
| Snellen Equivalent (mean, SD) |  |  |  |
| OD | 20/33 (32) | 20/32 (36) |  |
| OS | 20/26 (30) | 20/35 (35) |  |
| Location of Diabetes Diagnosis (%) |  |  | .3328 |
| During routine screening | 8 (72.7) | 31 (88.6) |  |
| During hospitalization | 3 (27.3) | 4 (11.4) |  |
| Prior dilated eye exam (%) | 8 (72.7) | 16 (45.7) | .1177 |
| Reported adherence to diabetic medication regimen (%) |  |  | .6395 |
| Somewhat | 2 (18.2) | 3 (8.6) |  |
| Very much | 9 (81.8) | 29 (82.8) |  |
| Unknown | 0 | 3 (8.6) |  |

<sup>a</sup> Three participants were excluded due to lack of gradable images.
<sup>b</sup> Point of care blood glucose (normal 4.0-5.4 mmol/L)
Abbreviations: SD, standard deviation; BMI, body mass index; LogMAR, Logarithm of the Minimum Angle of Resolution; OD, right eye; OS, left eye.
Continuous variables are presented with mean and standard deviation. Categorical variables are presented with N (%)

Out of the 95 eyes imaged, there was 100% agreement of ungradable images between graders and these images were excluded. For referral status, a third grader was required to reach a consensus in 11 eyes. There were 15 eyes (16%) that were considered to have referral-warranted diabetic retinopathy versus 69 eyes (73%) that did not require referral (Table 3). There was full agreement (κ = 1) on whether an image was of adequate quality for analysis. There was moderately high inter-rater reliability for referral status (κ = 0.62, 95% CI 0.42-0.83), retinopathy severity (κ = 0.76, 95% CI 0.55-0.96), and CSME (κ = 0.50, 95% CI 0.25-0.74; Table 4).

**Table 3.**
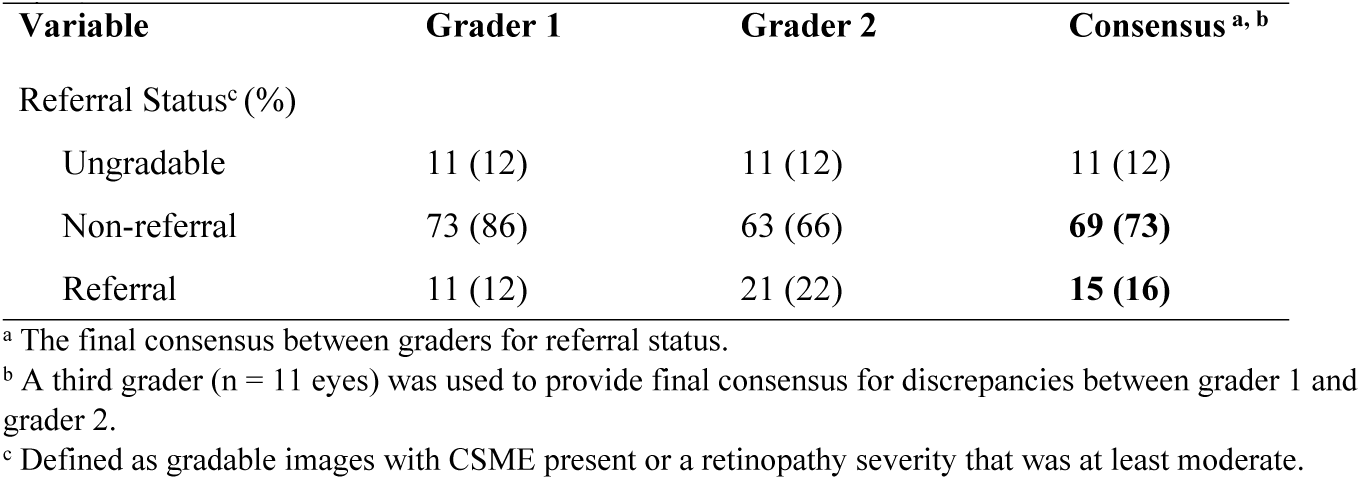
Referral status by grader based the diagnosis of DR in retinal images (n = 95 eyes)

**Table 4.**
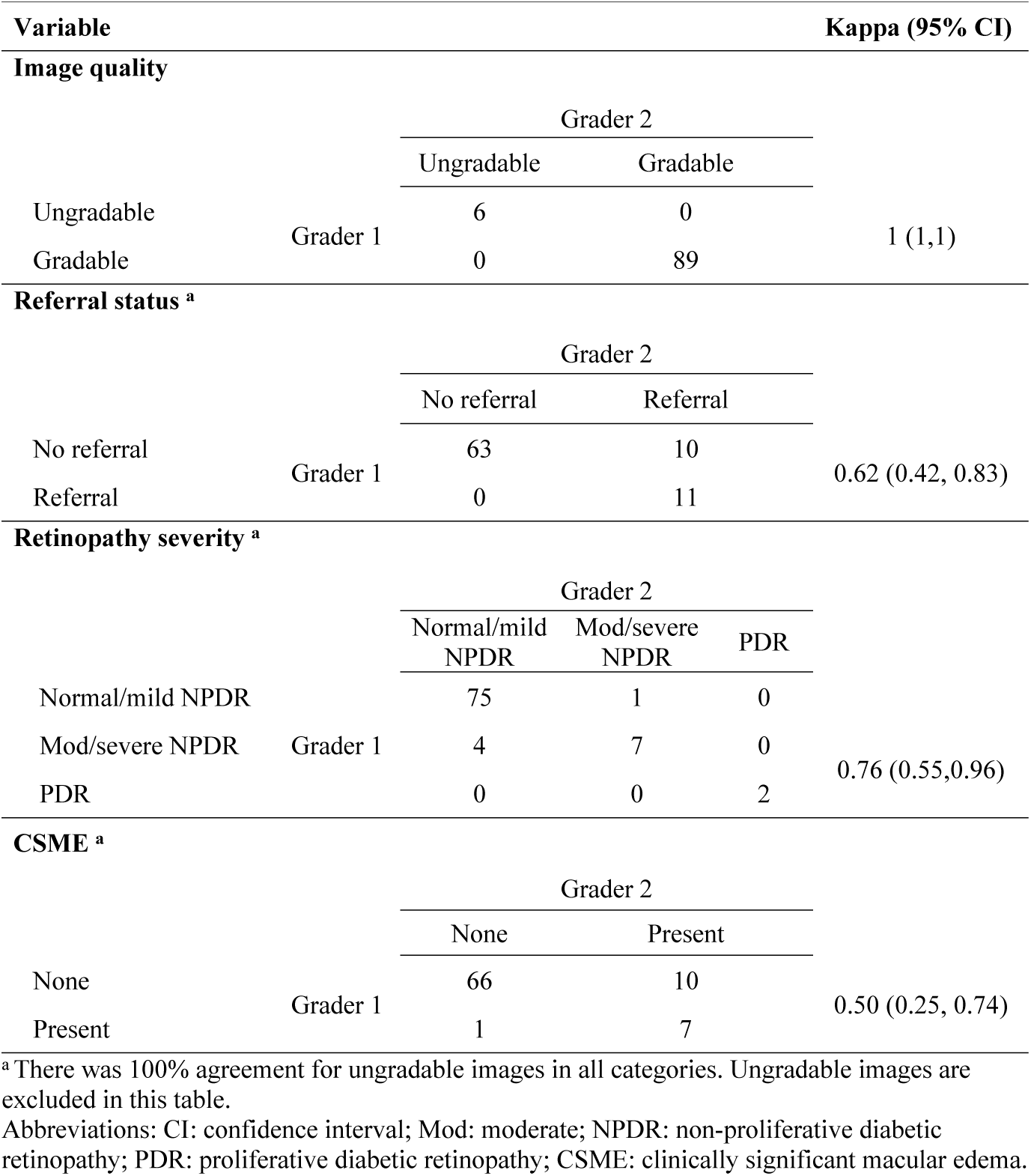
Inter-rater reliability between graders for image quality, retinopathy severity, the presence of macular edema, and referral status (n= 95 eyes)

## DISCUSSION

We performed a proof-of-concept study evaluating the use of a portable, handheld retinal fundus camera to screen for diabetic retinopathy in patients with diabetes in Vanuatu. We found that 24% of our cohort had referral-warranted diabetic retinopathy (DR), which was consistent with the 23.5% of participants with sight-threatening DR among diabetic patients in Vanuatu reported by Smith et al. in 2007(4). This high prevalence is problematic, especially due to the insufficient screening for DR in Vanuatu. Based on our data, only about half of the study population reported a prior dilated retinal exam, compared to almost 80% of U.S patients with diabetes who reported a dilated retinal exam over 24 months(22). This puts many of these patients at risk of vision loss and blindness due to inadequate early detection and intervention. The impact of pupil dilation was unclear, as most of our participants were dilated, although our main methodological goal was to obtain an adequate view of the fundus. The study did not evaluate the lens status. Nevertheless, there were multiple patients with cataracts that did limit the evaluation of the fundus. Furthermore, the elevated mean systolic blood pressure and blood sugar in this study indicate that these patients are at higher risk of developing DR(23). As such, an effective screening method for DR in Vanuatu is imperative.

The ideal screening modality in a low resource, rural setting must be cost-effective, portable, non-invasive, and reliable regardless of training level(6). We used the Volk Pictor Plus, a commercially available, handheld, non-mydriatic retinal camera. Accessible camera-based telemedicine screening programs have been shown to lead to saved sight-years(24). Although this camera costs approximately US $4,000-$10,000, this pales in comparison to the healthcare costs associated with blindness(25,26).

Due to the portability of the Pictor Plus, our team was able to travel across three different islands in Vanuatu without requiring extensive equipment. As for quality, only 12% of the retinal images taken were ungradable, despite the limited training of the photographers. This number of ungradable images is consistent with and sometimes less than prior studies screening for retinopathy in low-resource settings(27). We also found that about 16% of eyes (and 24% of participants) imaged had referral-warranted diabetic retinopathy, which is also consistent with the 17% of referral-warranted DR in a primary care study by Toy et al(28).

Furthermore, there was moderately high agreement between the two retina specialists in terms of referral status (κ = 0.62), retinopathy severity (κ = 0.76), and the presence of CSME (κ = 0.50). There was also full agreement (κ = 1) on whether an image was of adequate quality for analysis. For diabetic retinopathy screening purposes, images should be of adequate quality to be analyzed, which is why our analysis only differentiated between ungradable versus gradable images for quality. The images taken by our portable retinal camera were of enough quality to provide a reliable screening diagnosis of referral-warranted DR. While we did not evaluate the performance of the Volk Pictor Plus in this study, it has been shown to be 64-88% sensitive and 71-90% specific using a photographer with similar training(8). Although there is some controversy and variability in the literature about the quality of images taken with handheld non-mydriatic cameras, retinal photography can be obtained with high quality images with limited training(16,29,30). While most participants were dilated prior to image acquisition in this study, Sengupta et al. showed that there was only minimal improvement in sensitivity in patients with vision-threatening diabetic retinopathy when using a non-mydriatic camera without dilation. This would further facilitate image acquisition by photographers with limited experience(17).

The majority of the participants in the study (86%) were diagnosed with diabetes through screening held by the Ministry of Health in Vanuatu. These patients are subsequently followed at the non-communicable disease (NCD) clinic with routine BMI, blood pressure, and blood sugar measurements. Visual acuity measurement has also been implemented but is not regularly performed. While these variables are practical for assessing a patient’s disease progression, our study did not find them useful for predicting referral-warranted retinal disease, as there were no differences between patients with referral and non-referral-warranted disease. Nevertheless, if non-mydriatic portable retinal cameras are integrated in the NCD clinic workflow, they could be effectively used for screening in the primary care setting. Furthermore, local eye nurses could be effectively trained in the photographic detection of DR and identification of referral warranted DR as described by Boucher et al(31). Andonegui et al. similarly demonstrated that general practitioners can be trained to achieve high sensitivity and specificity in the detection of DR(32) with retinal photographs. By developing a tele-ophthalmology program, these retinal images can be stored and sent to the eye team in Port Vila and Santo for further analysis. This would allow for more frequent follow up for patients with severe DR, which should be done at least every 1-2 years in low-resource settings based on global guidelines(33). Artificial intelligence (AI) could also be used to perform image evaluation for detection of DR(34).

This proof-of-concept study was limited by multiple factors, although it does provide us with evidence that a portable retinal camera can be used for screening of diabetic retinopathy. The study’s small sample size could have led to a Type 2 error in the data analysis. In a larger, second-stage feasibility study, we may find a significant difference between groups. There was also a disparate representation of patients between the different islands studied, which could have biased the results. Given that participants were recruited to participate after their NCD clinic appointment, selection bias could have been introduced. However, this was the most efficient way to determine the prevalence of diabetic retinopathy in the diabetic population of Vanuatu in our proof-of-concept study. Retinal image quality was also limited for the purpose of prevalence analysis for numerous reasons. Photographers were not ophthalmic photographers and received limited initial training. However, this mimics the real-world limited training of photographers in many of these screening photography programs. Image quality could have also been compromised due to the language barrier between the photographers and the study participants. We were also unable to compare our images from images obtained from other imaging techniques, given the limited availability of imaging in Vanuatu. Nevertheless, our retina specialists used validated, consistent grading criteria and non-gradable images were not used in our analysis. The Pictor Plus has also been validated as a retinal camera in the studies mentioned above.

In summary, our study confirms the high prevalence of referral-warranted DR in Vanuatu. Our results suggest that handheld portable retinal cameras like the Volk Pictor Plus can be used to obtain retinal images of sufficient quality for effective diabetic retinopathy screening due to their portability, non-mydriatic capabilities, relatively limited training required for use, and cost-effectiveness. Moving forward, a validation study consisting of image acquisition by trained local nurses and image analysis by the Port Vila ophthalmology team could be performed to further evaluate the infrastructure capabilities for a telemedicine program in Vanuatu. A teleophthalmology screening program could assist with the prevention of blindness due to diabetic retinopathy in Vanuatu and other Pacific Islands.

## Data Availability

The data cannot be shared publicly because of HIPAA. Data is available from the University of Michigan Institutional Data Access/ Ethics Committee. Contact Dr. Paulus or Dr. Rosenthal for researchers who meet the criteria to access confidential de-identified data.

## AUTHOR CONTRIBUTIONS STATEMENT

Drs. Caceres, Zhang, Boe, Paulus, and Rosenthal made substantial contributions for the conception and design of the work. Drs. Caceres and Zhang made substantial contributions to the acquisition of the data. Drs. Caceres, Zhang, Besirli, Paulus, Rosenthal and Ms. Zhou made significant contributions to the analysis and interpretation of the data. All authors revised the work and approved the final submitted version. All authors also have agreed both to be personally accountable for the author’s own contributions and ensure that any questions related to the accuracy or integrity of the work are appropriately addressed.

## Acknowledgments

This work was sponsored by a grant from the National Eye Institute (1K08EY027458, PI YMP), Alcon Research Institute Young Investigator Grant (PI YMP), unrestricted departmental support from Research to Prevent Blindness, and the University of Michigan Department of Ophthalmology and Visual Sciences. We would like to thank the physicians, nurses, and staff at Godden Memorial Hospital, Northern Provincial Hospital, and Vila Central Hospital for all the support. We would also like to thank Alexis Cullen for helping us with the conception, design, and logistics of the trip. We finally thank Dr. David Musch for statistical guidance for the project.

